# The alternative complement pathway is activated in protoporphyria patients during sun exposure

**DOI:** 10.1101/2020.10.12.20210344

**Authors:** Francesca Granata, Lorena Duca, Valentina Brancaleoni, Silvia Fustinoni, Giacomo De Luca, Irene Motta, Giovanna Graziadei, Elena Di Pierro

**Author notes:** Corresponding author: Francesca Granata, MSc, Fondazione IRCCS Cà Granda Ospedale Maggiore Policlinico, Via F. Sforza 3520122 Milan, Italy.

## Abstract

The homeostasis of tissues in chronic disease is an important function of the alternative pathway (AP) of the complement system (CS). However, if not controlled, it may also be detrimental to healthy cells.

Protoporphyria (PP) is a rare disease that causes photosensitivity at the visible light due to the accumulation of Protoporphyrin-IX in the dermis. The aim of this study was to deep the knowledge about the involvement of AP in PP photoreaction.

Global radiation and UV data were provided from regional agency of environmental protection (ARPA). Properdin, Factor H (FH) and C5 levels were assessed in the serum collected during winter and summer from 19 PP patients and 13 controls..

Properdin in winter and summer reflected a positive increase compared to controls. The values in summer were higher than winter. The C5 results were altered only in summer. The outcome was reversed for FH: in the winter, it was higher compared to the summer. A positive correlation was reported between properdin and C3 in summer; a negative tendency between Factor B (FB) and FH was detected.

This study substantiated the differential involvement of AP depending on the increase in light exposure during the season, which was demonstrated with ARPA data. The enhanced systemic response could justify the malaise sensation of patients after long light exposure and can be exploited to elucidate the new therapeutic approach.

## INTRODUCTION

The complement system comprises of a network of 50 proteins and being a part of the innate immune system, it can be activated through three different pathways: classical, lectin, and alternative (also known as properdin pathway) (1).

Several investigations over the last decade tried to decipher the function of the alternative pathway (AP) in inflammation (2), especially in chronic disease (arthritis, asthma, and kidney failure). Active participation of this pathway has been well established in regulating the homeostasis of tissue, eradicating cellular debris, orchestrating immune responses, and sending “danger” signals (3-5). However, if the alternative pathway of the complement system becomes uncontrolled, it can also attack the healthy cells resulting in tissue destruction and aggravation of symptoms in the acute phase of some diseases (stroke, atypical hemolytic uremic syndrome (aHUS) or primary thrombotic microangiopathy (TMA) (6-9). Properdin, a multimeric protein, takes part in the activation of the AP by acting as a positive regulator of the amplification loop (10). Properdin possesses a strong binding affinity for C3b, originating from the spontaneous hydrolysis of C3 protein in the fluid phase (Figure 5) (11, 12). Under physiological conditions, the hydrolyzed C3 has a short half-life of ∼90 seconds both in the fluid phase and on cell surfaces (13). The C3 and C5 convertases complex are stabilized on binding with properdin and the Factor B (other protein of the AP) that increase their half-life by 5–10-fold (2). This mechanism leads to the activation of the AP loop answer. Another important phenomenon involved in the regulation of AP is the Factor H competing with FB for binding to C3b, in turn, inhibiting the assembly of the C3 convertase and facilitating the degradation of the already formed complex (14). The protective mechanism thus functions as a sentry to ensure that only when it is required, the spontaneous activation of C3 is stabilized by properdin and factor B with a consequent stabilization of C3b subunit and activation of C5 (3).

**Figure 1.**
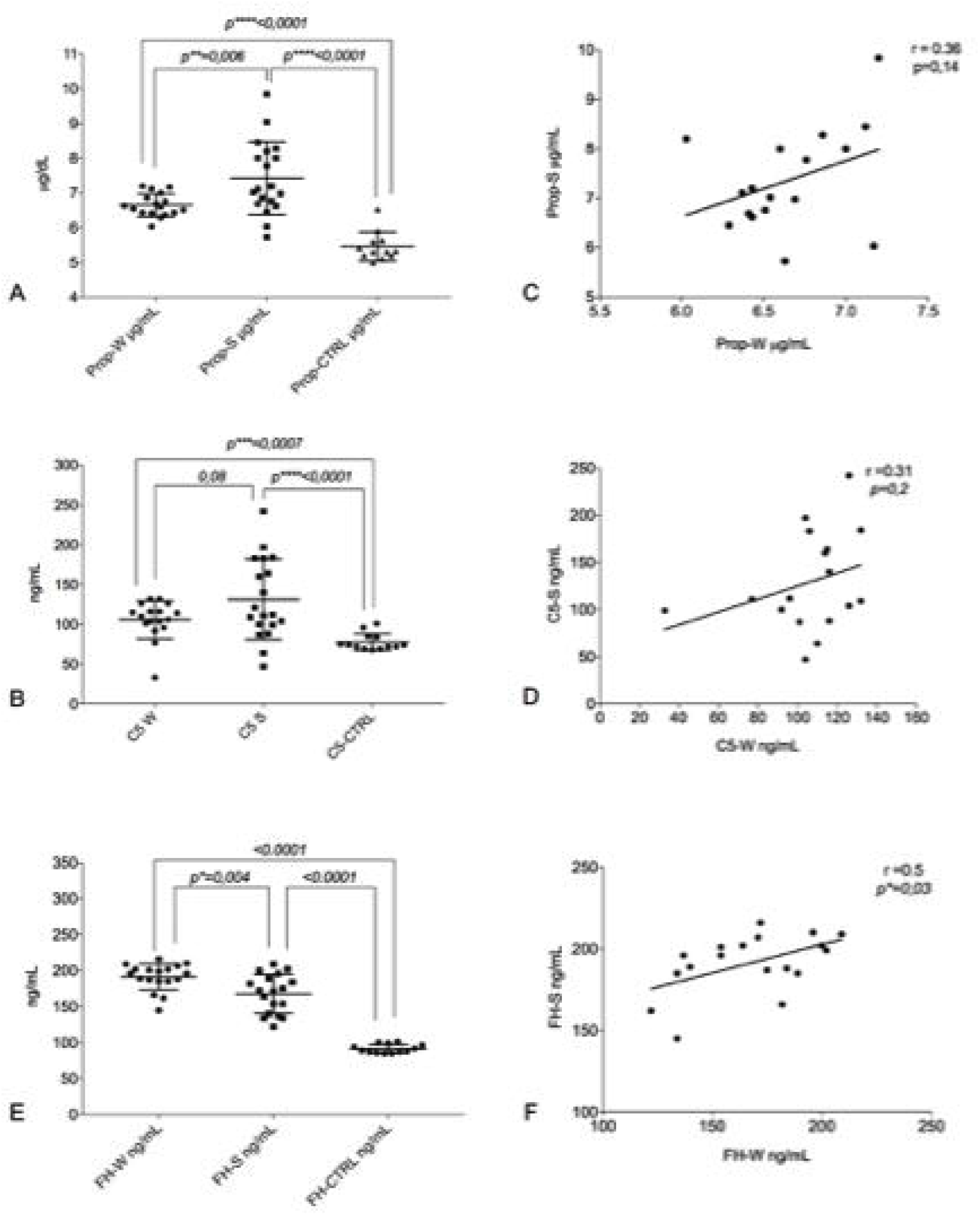
Protein of AP assays. ***a***. Distribution of the mean levels and ±DS of properdin in winter (Prop-W µ = 6.6 ±0.33); properdin in summer (Prop-S µ = 7.4 ±1.05) and the control group (Prop-CTRL µ = 5.4 ±0.42 ***b***. Distribution of the mean levels and ±DS of C5 in winter (C5-W µ = 105.9 ±23.7); C5 in summer (C5-S µ = 131–3 ±50) and the control group (C5-CTRL µ = 77.5 ±10.6) ***c*** Correlation between the seasonal values in PP patients of properdin (r = 0.36 p = 0.14) ***d*** Correlation between the seasonal values in PP patients of C5 (r = 0.31, p = 0.2). Factor H (FH) assay. ***e***. Distribution of the mean levels and ±DS of FH in winter (FH-W µ = 191.4 ±18.31); FH in summer (FH-S µ = 167.7 ±26) and the control group (FH-CTRL µ = 91,4 ±15.9). ***f***. Correlation between the seasonal values in PP patients for FH (r = 0.5, p*= 0.03).

**Figure 2.**
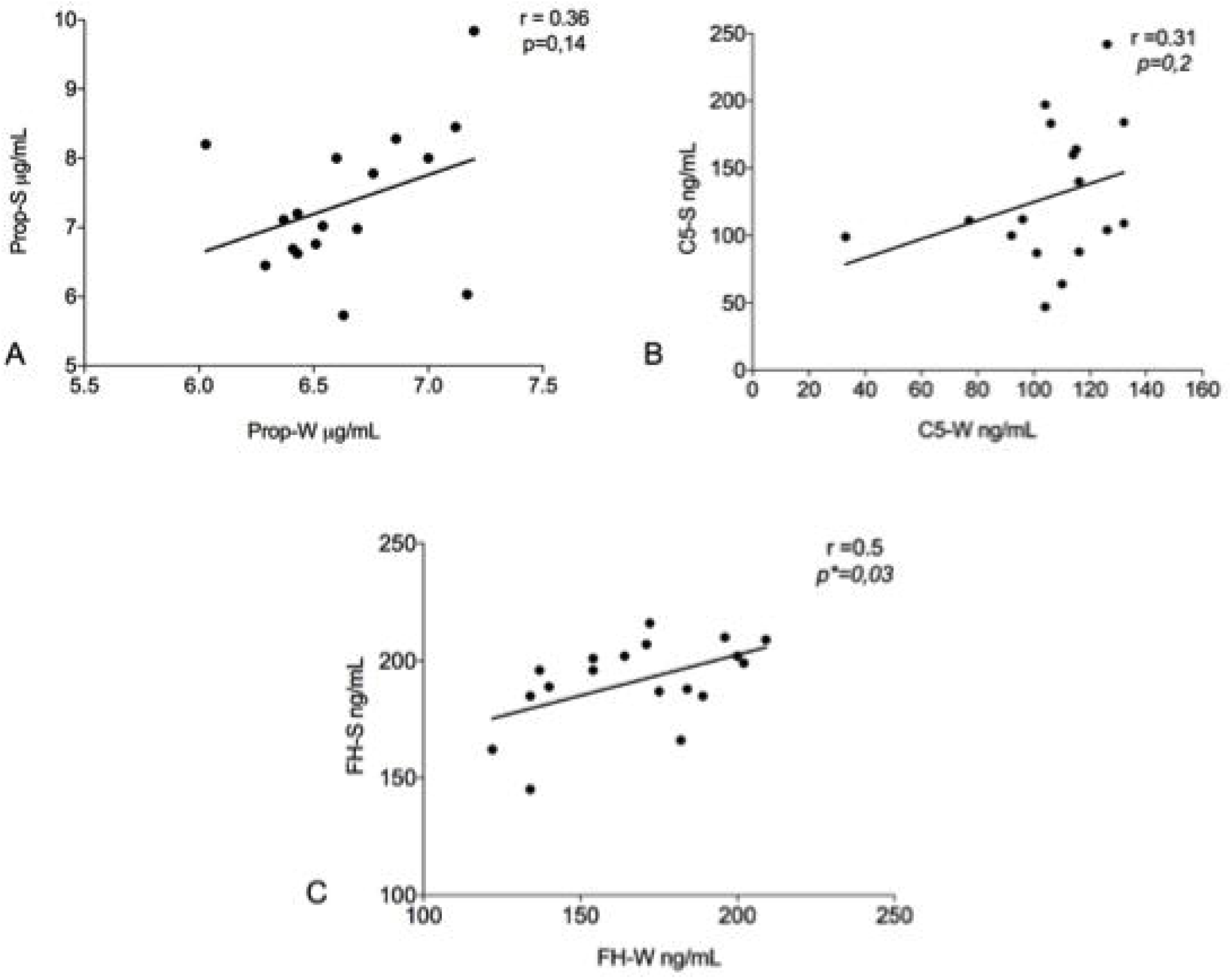
Correlation between the results of the alternative pathway proteins in summer. a. Positive correlation between properdin and C3 in winter (r = 0.49 p =**0.03) b. Positive correlation between properdin and C3 in summer (r = 0.71 p =***0.005), c. Negative tendency between FH-S and FB-S data previously show by Granata et al. 2019 (r = − 0.42 p = 0.07).

**Figure 3:**
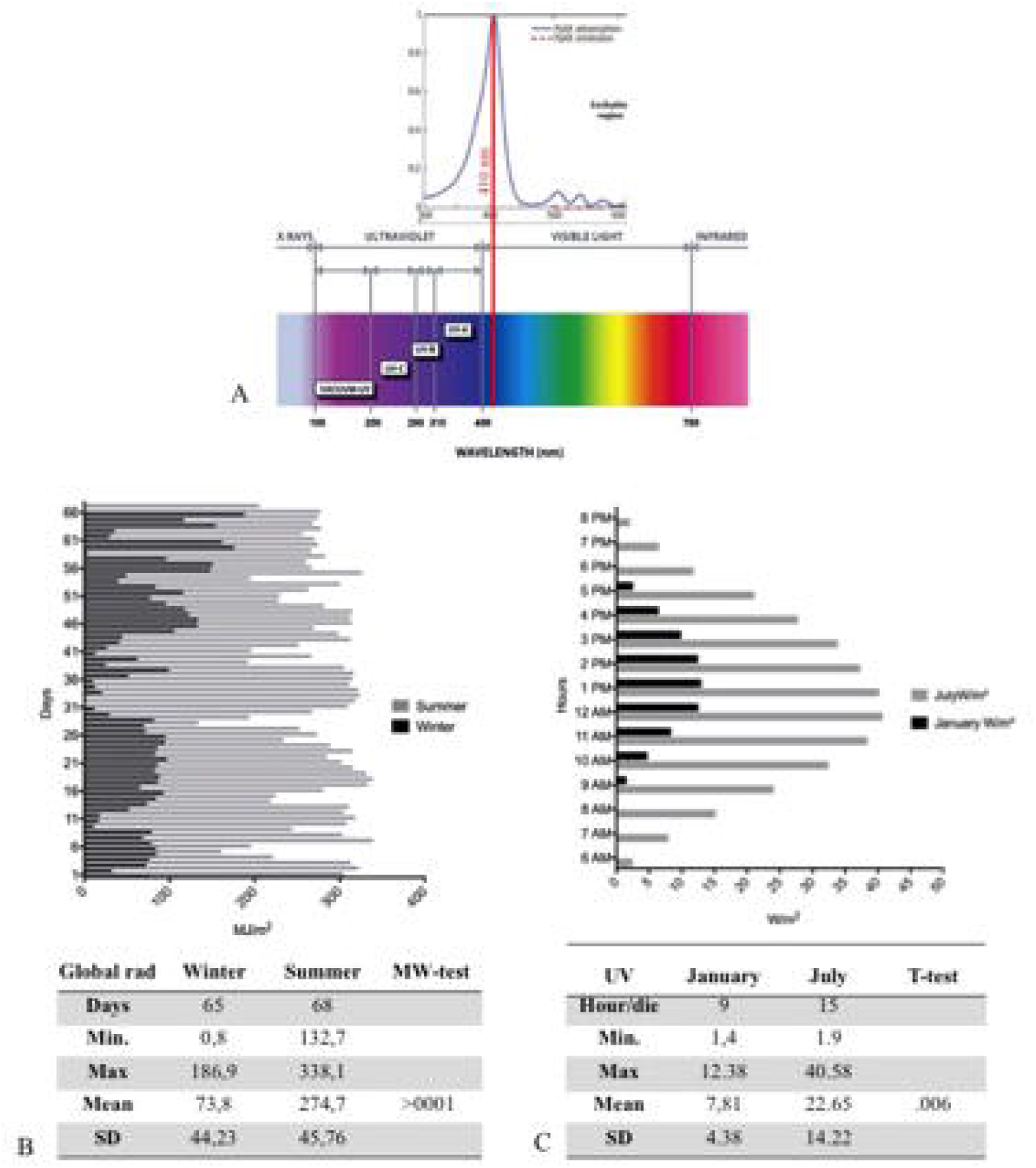
The Global Radiation, UV radiation and PPIX peak. a. Upper: The portion of the PpIX absorption spectrum with a maximum peak around 410 nm in visible. Lower the global radiation spectrum with different wavelength UVC (200-280nm) UVB (280-315nm) UVA (315- ≈ 400 nm) and visible light (≈400-780). b. The daily mean detection by ARPA of global radiation in 65 days in winter (black) and 68 days of summer (gray). The figure shows a high summer trend of global radiation compared to winter. In the figure are sum-up the statistical values with a significant mean difference (>0001) between seasons. c. The Daily hours in January and in July, during the maximal variability of UV ray. In the figure are sum-up the statistical values with a significant mean difference (>006) between seasons.

**Figure 4:**
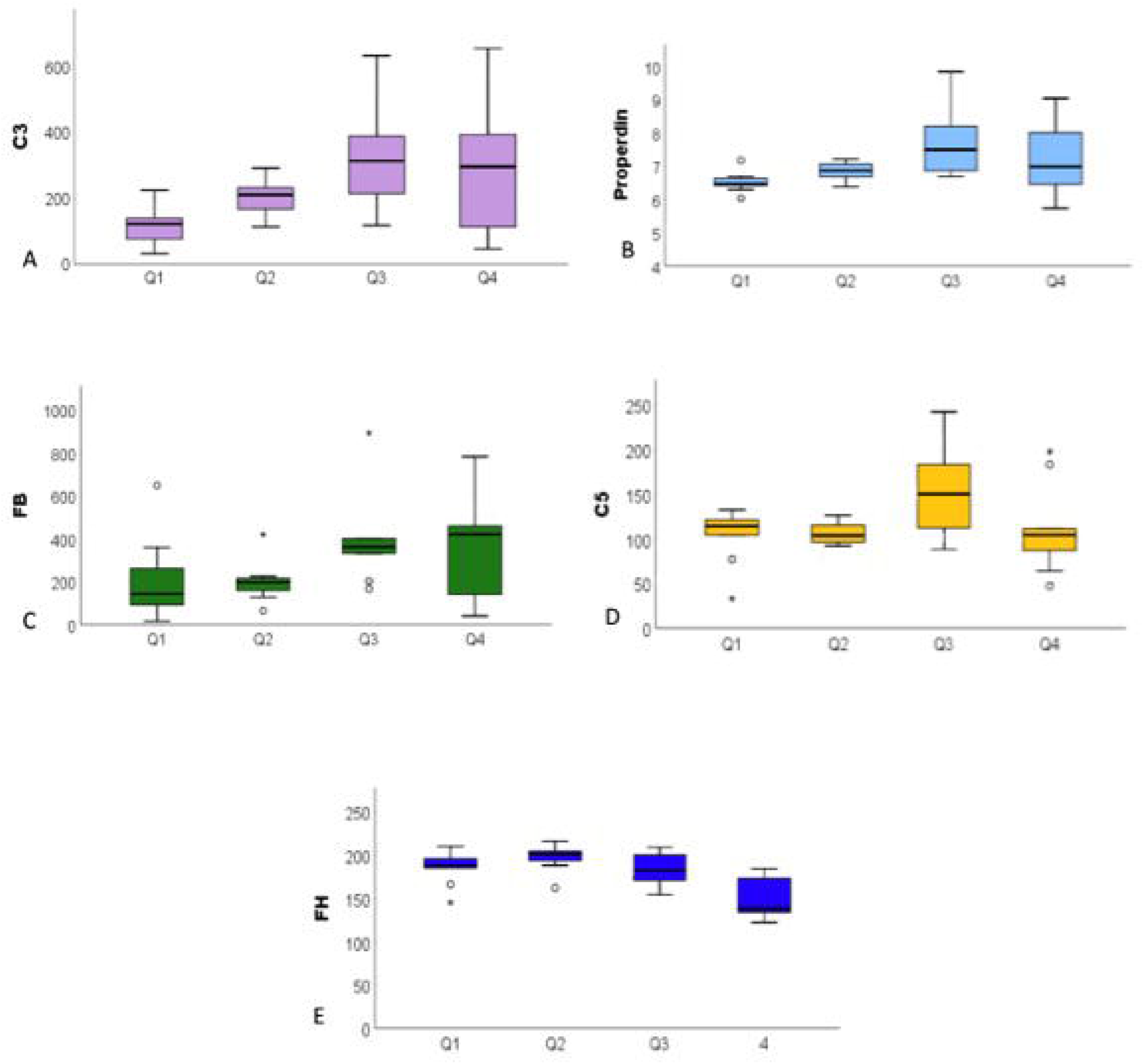
The ANOVA analysis among the exposure to global radiation in the 14 days before blood sampling related to complement system proteins. a. Significative increase of C3 among the quartile (<007), in particular to Q1 from Q3, with a stabilization in Q4 b. properdin significate increasing (<0.016) c. Increased FB values among quartiles (<0.08) d. stable distribution of C5 (<057) e. FH decreasing among quartile (<0.001)

**Figure 5:**
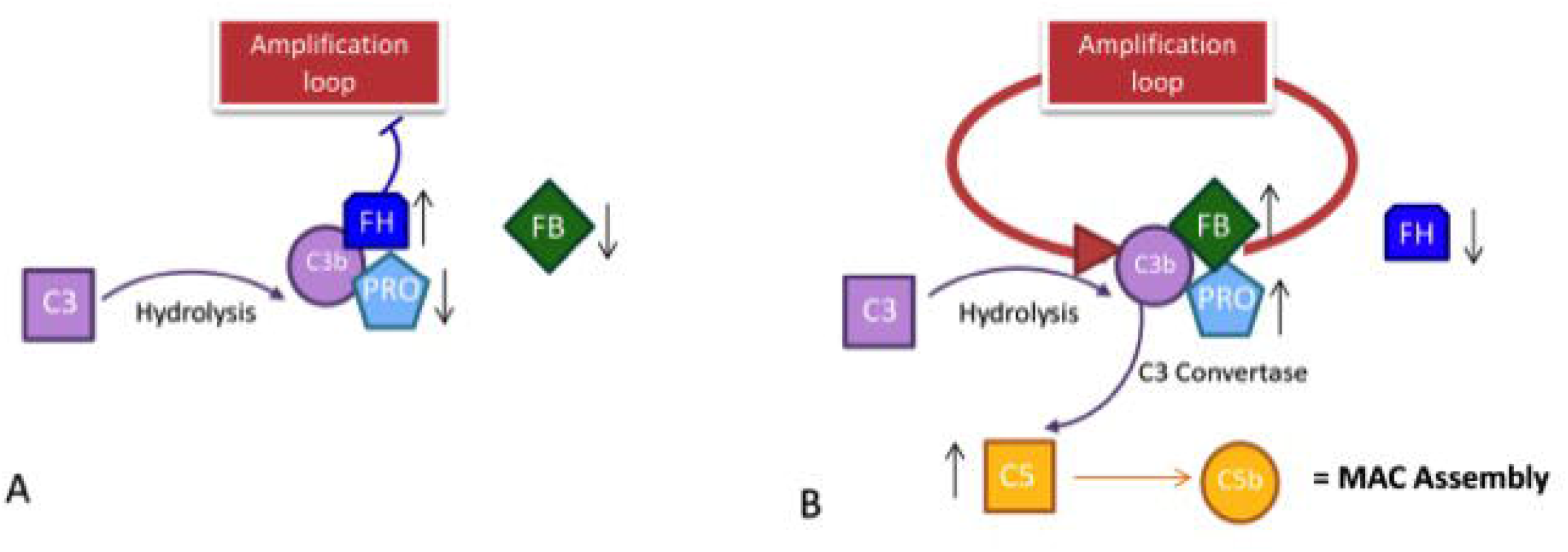
The course of the alternative pathway in PP patients during the season. A. **In winter**, the decrease in solar irradiation does not trigger the establishment of an amplification loop, because the level of properdin and FB is low. The inhibitor factor H in winter must be high to occupy the C3b site for the spontaneous hydrolysis of C3; this prevents the stabilization of C3 convertase and activates the loop amplification mechanism. B. **In summer**, the data show the establishment of an amplification loop; the spontaneous hydrolysis of C3 in C3b is stabilized by the increase in properdin and Factor-B that stabilize the C3b in C3 convertase complex. In summer, the level of inhibitor FH is decreased compared to winter because the link with C3b is already occupied by FB by which it competes. C3b activation leads to the activation of C5 with consequent MAC assembly activation.

Erythropoietic protoporphyria (EPP; OMIM 177000) and X-linked protoporphyria (XLP; MIM 300752) are two rare disorders, triggered respectively by the deficiency of the enzyme ferrochelatase (FECH EC 4.99.1.1) or the activation of the erythroid specific form of 5-aminolevulinate synthase 2 (ALAS2; EC 2.3.1.37) in the heme biosynthesis pathway (15)[(16). The exacerbation of the symptoms is due to the accumulation of protoporphyrin-IX (PP-IX) in erythrocytes and tissues, in particular in dermis and consists in severe photo reaction. The PPIX was excited at 410 nm in the Soret Band of visible light, with significant emission peaks of activation at 635 nm (figure 3A) (17). The visible spectrum partially overlaps the UV-A (from ∼ 360–380 to 400 nm in the deep violet) ending at 780nm. (figure 3A) (18). Thus, the PPIX peak shows its ascending curve in this region between late UVA and visible spectrum, with its maximum peak at 410 nm. The visible light of global solar radiation is not limited to the wavelengths in the visible, but it includes the total short-wave radiation falling from the sky onto a horizontal surface on the ground. Also, it includes both the direct solar radiation and the diffuse radiation resulting from reflected or scattered sunlight (19).

Within just few minutes of exposure to this wavelength, individuals with this disease develop extremely painful cutaneous conditions and the symptoms include burning, acute skin pain, itching, and edema (20). The biological process that leads to a phototoxic reaction in patients with protoporphyria has not yet completely elucidated. It is widely known that the phototoxic reaction occurs because of the increased level of reactive oxygen species (ROS) in the derma that leads to endothelial cell photo-damage (21) through complement system activation and mast cell degranulation, culminating into exocytosis of vasoactive mediators and acute inflammation (22).

The detection of the complement system proteins, C3 and C5, in blood samples of only two PP patients, collected after skin irradiation (0.7 J/cm2 at 400–410 nm), confirmed the involvement of the complement system in the pathophysiology of PP symptoms (23). Our recent study, conducted through the assessment of two specific proteins, Factor B for the AP and C1q from the classical pathway, excluded the involvement of these latter, through the detection of the normal level in both seasons of C1q protein. Enhanced levels of FB in summer suggested the involvement of the alternative pathway (AP) in the molecular mechanisms leading to the phototoxic reaction (24).

Considering the previously obtained data on Factor B and C3, the present study aimed to estimate in detail the activation of the alternative pathway through the assessment of the properdin, factor H, and C5 for in-depth knowledge of phototoxic reaction in PP patients. Furthermore, through the collection of global radiation data, we have estimated the personal exposure to global radiation in PP patients, in order to better investigate the relationship between this exposure and the metabolites of the AP.

## MATERIAL AND METHODS

### Patients

The study involved a total of 19 PP patients diagnosed and referred to Rare Diseases Centre at Foundation IRCCS Ca’ Granda Policlinico of Milan and 13 healthy individuals aged-matched and randomly collected during the seasons. To verify the reaction to the light exposure in each group, with any further invasive treatment, blood samples were collected during routine test at two points in winter (January, February and first week of March 2017) and summer (June, July, or the first week of August 2017), respectively. Routine blood panel analyses include specific porphyria parameters as erythrocyte-free protoporphyrin-IX (PPIX); zinc proto-porphyrin (ZPP) and plasma peak performed following the methods previously described (25). According to the World Medical Association Declaration of Helsinki for medical research, all subjects involved in this study signed informed consent for the diagnosis and research approved by the ethics committee of our institution, Fondazione IRCCS Ca’ Granda–Ospedale Maggiore Policlinico and the identity of the study participants were anonymized.

### Laboratory testing for C5, Properdin and Factor H

Serum was separated from whole blood by centrifuging at 10,000 rpm at 4°C for 10 min and stored at –80 °C for future use. Serum C5, Properdin, and Factor H were determined using a commercial immunoassay (Hycult Biotech) kit following the manufacturer indications (Hycult Biotech Uden, The Netherlands). The two different samples from the PP patients were used to examine each marker, and the results were compared to values registered in healthy subjects

### Global radiation (GR) data and UV data

The service ‘IdroNivoMeteo e Clima’ of Agenzia Regionale Protezione Ambientale–ARPA Lombardia provided the sun global radiation data from January 2017 to August of 2017 for the Lombardy area. The data were transmitted as daily average radiation (MJ/m^2^). Moreover, the raw data of UVA (W/m^**2**^) expressed in hour per day, from the different weather stations in Lombardy, expressed in W/m^**2**^, were given. The mean radiation received by each patient in the 14 days preceding the blood sampling was calculated and used as a surrogate of the personal exposure to global radiation (MJ/m^2^).

### Statistical analysis

The data analysis was executed using GraphPad Prism software version 7. D’Agostino–Pearson’s normality test, Shapiro–Wilk normality test, and the KS normality test were applied to confirm the normality of the data from each measurement. In order to identify the outliers, the ROUT (1%) test was performed for each experiment. Unpaired or paired parametric t-tests and Mann-Whitney U-test were employed for statistical analyses between seasons (winter vs. summer) and controls. Pearson’s correlations were determined between two datasets with a two-tailed and confidence interval at 95%. Linear regression was also calculated at a confidence interval of 95%. Box-plots were used to represent the distribution of biological parameters at different quartiles of personal exposure to global radiation; ANOVA was used to compare the groups.

## RESULTS

### Patients’ characteristics

The clinical and biochemical findings for individuals involved in the study are summarized in Tables 1. Nine males, aged 39 ±10.4 years and 10 females, 39±10.3 years with PP were involved in this study. They exhibited typical plasma fluorescence peak at 631-635 nm. Total erythrocyte protoporphyrin-IX (with the percentage of proto/zinc-PP) was increased in all patients, no differences were observed between males and females (respectively µ = 77 ±38 and µ = 76 ±37). FECH mutation was described for every patient and reported in table 1.

**Table 1:** Clinical and biochemical findings.

| Patients | Age | Sex | Peak | PP totali<br>(mg/gHb) | Proto<br>% | Zinco<br>% | FECH analysis |
| --- | --- | --- | --- | --- | --- | --- | --- |
| Pt1 | 36 | M | 632 | 123,7 | 97 | 3 | FECH: c.[1-251G>C;194+4350_463+1197del5577];[315-48T>C] |
| Pt2 | 32 | F | 634 | 63,4 | 91 | 9 | FECH: c.[215dupT];[315-48T>C] |
| Pt3 | 40 | F | 634 | 84,2 | 39 | 0 | ALAS2: c.[1706_1709delAGTG];[=] |
| Pt4 | 29 | M | 634 | 90,4 | 96 | 4 | FECH: c.[1-251G>C;194+4350_463+1197del5577];[315-48T>C] |
| Pt5 | 36 | M | 633 | 64,4 | 98 | 2 | FECH: c.[215dupT];[315-48T>C] |
| Pt6 | 45 | F | 634 | 63,9 | 95 | 5 | FECH: c.[901_902delTG];[315-48T>C] |
| Pt7 | 51 | F | 633 | 165,4 | 97 | 3 | FECH: c.[901_902delTG];[315-48T>C] |
| Pt8 | 36 | F | 635 | 94,2 | 95 | 5 | FECH: c.[215dupT];[315-48T>C] |
| Pt9 | 51 | M | 634 | 67 | 96 | 4 | FECH: c.[464-1169 A>C];[315-48T>C] |
| Pt10 | 25 | M | 632 | 38 | 94 | 6 | FECH: c.[215dupT];[315-48T>C] |
| Pt11 | 27 | F | 631 | 41 | 92 | 8 | FECH: c.[215dupT];[315-48T>C] |
| Pt12 | 54 | F | 634 | 55 | 96 | 4 | FECH: c.[464-1169 A>C];[315-48T>C] |
| Pt13 | 24 | M | 634 | 63,7 | 96 | 4 | FECH: c.[67+5G>A];[315-48T>C] |
| Pt14 | 40 | M | 634 | 95,7 | 98 | 2 | FECH: c.[215dupT];[315-48T>C] |
| Pt15 | 27 | F | 631 | 61,8 | 94 | 6 | FECH: c.[1080_1081delTG];[315-48T>C] |
| Pt16 | 49 | F | 634 | 29,6 | 93 | 7 | FECH: c.[215dupT];[315-48T>C] |
| Pt17 | 48 | M | 631 | 33,2 | 92 | 8 | FECH: c.[464-1169 A>C];[315-48T>C] |
| Pt18 | 54 | M | 635 | 152,4 | 98 | 2 | FECH: c.[215dupT];[315-48T>C] |
| Pt19 | 37 | F | 634 | 55,7 | 96 | 4 | FECH: c.[1-251G>C;194+4350_463+1197del5577];[315-48T>C] |

### Increase of Properdin and C5 levels and decrease of inhibitor factor-H in PP samples between seasons

Compared to the healthy subjects (Prop-CTRL), marked difference (< 0.0001) in properdin levels of PP patients both in winter (Prop-W) and in summer (Prop-S) were noted. Interestingly, the comparison between Prop-W and Prop-S also revealed showed a significant increase in properdin during summer (< 0.006) (Fig 1A). A statistically significant difference was also observed between C5-S levels in winter (C5-W) and summer (C5-S) in PP patients compared to the controls (p < 0.007 and p = 0.0001, respectively). A slight increase in C5 was also evident when comparing summer (C5-S) and winter (C5-W) values in PP patients (p < 0.08) (figure 1B). We also observed a positive correlation tendency between properdin and C5 in both summer and winter (respectively, r=0.36, p=0.14, r=0.31, p=0.2 and) (figure 1C and 1D).

The mean value of factor H in winter (FH-W) was higher mean compared to the summer (FH-S) in PP subjects (p=0.004). Moreover, PP patients’ values either from summer or winter were significantly higher compared to healthy controls (FH-CTRL) (< 0.0001) (Fig 1E).

The escalation of FH between seasons (winter vs. summer) was associated with a positive intra-patient correlation (r=0.5, p=0.03) (Fig 1F).

### Correlation between the major important proteins of the alternative pathway

Following the same methodology, data about C3 and Factor B were previously obtained from the same samples (24). Here, we report an additional analysis with data obtained in this study. A significant positive correlation (r =0.49, p=0.03) between C3 and properdin values in winter PP samples was established (Fig 2A). Figure 2B represents the correlation between the same factors in summer, which showed an even stronger positive correlation (r=0.71, p=0.005). On the contrary, a negative correlation was seen between FB-S and FH, i.e., FH increased with a gradual reduction in FB (r = –0.42, p = 0.07) (figure 2C).

### Personal exposure to global radiation and its relationship with the complement metabolites

Figure 3A represents the radiation spectrum with the PpIX absorption lambda to 410 nm in visible. Figure 3 B shows different trend of global radiation between the seasons, in winter it was registered for 65 days with a mean and SD of µ = 73,8 ±44,23, that was statistically different compared to 68 days of summer detection (µ = 247,7 ± 45,76) (p <0.001). Moreover, figure 3C represent the mean and SD of UVA by hour per day in January (µ = 7,81 ± 4,83) and July (µ = 22,65 ± 22,41).

The global radiation collected from ARPA during the 14 days prior to the blood draw was divided into quartiles: Q1 (51.3-65.7 MJ/m2); Q2 (75.4-148.6 MJ/m2); Q3 (159.5-283.1 MJ/m2); Q4 (286.0-297.9 MJ/m2). The distribution of metabolites of the complement system in the different quartiles of global radiation exposure was depicted using box-plots. Comparing the groups, we observe significant or marginally significant differences for the alternative pathway metabolites: C3 (p = 0.007); properdin (p = 0.016); FB (p = 0.084); C5 (p = 0.057); and FH (p <0.001). C3 and properdin showed positive increase when the exposure to global radiation, while FH showed a decreasing trend. The C5 and FB show marginally significant differences (Figure 4A-E).

## DISCUSSION

The phototoxic reaction is one of the predominant clinical symptoms in PP, our work highlights that reaction is not only attributable to a local response in the dermis of the patients but include a systematic involvement. The symptoms experienced by some patients and described as chills, malaise, fatigue; nausea; excessive temperature sensitivity or generally unwell after long solar exposure (26, 27), could be attributed to the implication of the immune system.

A combination of multifactorial conditions including seasons, cloud cover, intensity and extent of sun-exposure, and time of day (28) is responsible for the PP symptoms heterogeneity, whose intensity varies from mild to severe. Furthermore, the patients are aware of the limit beyond which even mild symptoms can occur (29). Therefore, the study of a systemic involvement cannot take place with intensive exposure to sunlight. Our method has allowed us to analyze the complement system fluctuations associated with the variation of light intensity between seasons without exposing the patients to dangerous and painful treatments. In our previous work we reported a significant increase in the C3 and FB proteins, primarily during the summer season, which highlighted the involvement of the alternative pathway of the complement system in the phototoxic reaction of PP patients (24).

Here, we report additional experiments that strengthen this presumption. The properdin is the main protein that binds the FB-C3b complex and it triggers the amplification loop in the alternative pathway (Figure 5B). Properdin was found to be positively correlated with the C3 in summer PP samples (figure 2B) suggesting the AP activation during this season.

However, PP sample in winter (compared to control) showed enhanced levels of properdin too, suggesting the stimulation of AP also during this season. The exposure of the patients to global radiation also during in winter (figure 1B) could justify the above findings. Moreover few patients may also be sensitive to some types of artificial light that triggers the reaction (30).

Furthermore, being the increase of FB proportional to the decrease of FH, it further supports the theory of the strong and looping answer of AP to light exposure in summer.

The lower levels of FH in summer compared to winter should cause a loss of control over the loop of AP corroborating the thesis of the AP activation. FB competes with FH for the same C3b binding site and its increase proportional to the FH decrease in PP samples. Taken altogether this supports the theory of a strong and looping answer of AP to light exposure in summer.

The higher level of C5 proteins in summer than winter, resulting in the formation of the membrane attack complex (MAC) assembly for cell lysis in summer, also substantiated this evidence.

A differential function of the complement system has been already reported in other chronic or acute pathologic conditions (3-5). Thus similarly also in PP, the excitation of PPIX should activate both functions of the AP complement system compared to healthy controls. In winter, the intensity and duration of global radiation (Figure 3b-3c) and less sun-exposed skin areas should lightly stimulate PPIX causing only a mild activation of the complement system (up regulation of FB; Properdin; C5; FH). This should represent the chronic branch of system, which can in turn favor the tissue homeostasis and maintain the patient in a free symptoms status. Therefore AP can respond to the signal for tissue repair and recovery with minimal cell damage in winter (chronic phase) and accordingly acts as the mechanism of alert (12).

On the contrary, in summer a stronger intensity and longer sun radiation with bigger sun-exposed skin areas could acutely activate the AP loop. The enhanced light intensity and exposure in summer causes a rapid increase of AP proteins and a decrease of the inhibitor factor H. The loss of inhibition activates the AP loop and an acute response, resulting in tissue destruction and aggravation of symptoms that can last up to ten days (31). The length of symptoms could be better justified by the involvement of systemic answer and they could not be only due to the time of decay of excited PPIX. The decay of excited PPIX is quite fast; the visible light absorption creates a singlet-excited state in PPIX (1PP*), with nanosecond lifetime, which undergoes a fast relaxation to excited triplet state (3PP*) with duration of milliseconds. During this time the PPIX transfer its energy to molecular oxygen (O2) with a consequent decay of excited PPIX. Therefore the long lifetime of symptoms could be better justified by the implication of systemic answer and not be only due to the time of decay of excited PPIX. (32)

All these assumptions are also supported by the evaluation of exposure to global solar radiation. As the global radiation increase, the main factors of the alternative pathway such as C3 and properdin increase. The FH values divided into quartiles decrease with increased of global radiation in Q3 and Q4, this confirms the loss of inhibition of activation of the AP loop in summer. The protein C5 and FB showed marginally significant differences but with an increasing tendency throughout the quartiles. This uneven growth could be explained by the fact that the method is not designed on patients but represents a collection of global radiation values during the 14 days prior to the blood drawn. During this period some patients may not have been fully exposed. It is well known that PP patients change their behavior during the summer season to avoid sun exposure and the maximum of the symptoms (33) and consequently avoiding the activation of the loop. In particular, the C5 represents the last step of activation, before the MAC assembly and apoptosis of damaged cells and that PP patients, at the time of the summer sample collection, were not in the acute phototoxic state.

To improve the quality of life of the patients for several diseases, management of symptoms and pain is of utmost importance (34, 35). Likewise, for PP, a constant challenge prevails in daily life to avoid the pain that leads to reduced quality of life (36). Moreover, analgesic drugs fail to relieve the pain in PP (31).

In conclusion, the new insights provided by this study about the involvement of the systemic response in PP can help to understand anti-inflammatory mechanisms of existing drugs as alpha-melanocyte–stimulating hormone (α-MSH) or identifying newer therapeutic targets, able to interrupting the systemic response in EPP or useful for pain management in photodynamic therapy (PDT) (32, 37).

Future study will be necessary for personalizing the detection method of visible light exposure. At the same time, also a more suitable *in vitro* setting to evaluate the level of AP activation during different timelines of light exposure should be developed, thus avoiding the involvement of the patient in invasive treatment. This will allow to gain a deeper insights of the clinical symptoms complained by the patients, such as the malaise after a long sun exposure.

## Data Availability

The authors declare that they have no conflict of interest.

## Informed consent

Informed consent for the research was obtained from all individual participants included in the study.

## Author contributor

FG designed the study, performed statistical analysis, and wrote the manuscript; LD performed the ELISA experiments; VB carried out a genetic diagnosis of patients; SF manager of toxicology lab of Policlinico and suggest the method for global radiation and UV detection from ARPA data; GDL recruited the patients; IM oversaw the work; GG recruited the patients; EDP critically revised the results and manuscript. The manuscript has been read and approved for submission by all the named authors.

## Acknowledgments

We are indebted to all patients who participated in this research and to the DH staff of UOC rare centre diseases at Fondazione IRCCS Ospedale Maggiore Policlinico, who assisted every day the PP patients, especially during the summer season. The author would like to acknowledge, with gratitude, MD Maria Itala Baldini, for her constant support. We would like to thanks Maria Domenica Cappellini to give us the possibility of work in this field. We would like to thanks to ARPA the IdroNivoMeteo service of Regione Lombardia who provides us the global radiationand and UV data for our work.

